# Rapid classification and prediction of COVID-19 severity by MALDI-TOF mass spectrometry analysis of serum peptidome

**DOI:** 10.1101/2020.10.30.20223057

**Authors:** Rosa M. Gomila, Gabriel Martorell, Pablo A. Fraile-Ribot, Antonio Doménech-Sánchez, Antonio Oliver, Mercedes García-Gasalla, Sebastián Albertí

## Abstract

Classification and early detection of severe COVID-19 patients is urgently required to establish an effective treatment. Here, we tested the utility of matrix-assisted laser desorption/ionization time-of-flight mass spectrometry (MALDI-TOF MS) to classify and predict the severity of COVID-19 in a clinical setting. We used this technology to analyse the mass spectra profiles of the sera from 80 COVID-19 patients, clinically classified as mild (33), severe (26) and critical (21), and 20 healthy controls. We found a clear variability of the serum peptidome profile depending on COVID-19 severity. Seventy-eight peaks were significantly different and 12 at least four fold more intense in the set of critical patients than in the mild ones. Analysis of the resulting matrix of peak intensities by machine learning approaches classified severe (severe and critical) and non-severe (mild) patients with a 90% of accuracy. Furthermore, machine learning predicted correctly the favourable outcome of the severe patients in 85% of the cases and the unfavourable in 38% of the cases. Finally, liquid chromatography mass spectrometry analysis of sera identified five proteins that were significantly upregulated in the critical patients. They included serum amyloid proteins A1 and A2, which probably yielded the most intense peaks with *m/z* 11,530 and 11,686 detected by MALDI-TOF MS.

In summary, we demonstrated the potential of the MALDI-TOF MS as a bench to bedside technology to aid clinicians in their decisions to classify COVID-19 patients and predict their evolution.

## INTRODUCTION

Coronavirus infectious disease 19 (COVID-19) was first reported in Wuhan, Hubei province, China as a new coronavirus disease caused by a positive-strand RNA virus designated as severe acute respiratory syndrome coronavirus 2 (SARS-CoV-2) (1). This virus is responsible of a pandemic of unprecedented dimensions. Approximately 80% of COVID-19 cases are asymptomatic or present mild symptoms, such as fever, cough, fatigue, and dyspnea. Conversely, about 20% of patients with COVID-19 develop viral pneumonia, with an exaggerated host inflammatory response and hypoxia, requiring intubation and mechanical ventilation (2,3). These patients, classified as clinically severe or critical life threatening infections, are mainly diagnosed empirically based on a set of clinical characteristics. However, patients with these symptoms have already evolved to a serious clinical condition that requires specialized intensive care. Therefore it is essential to set up novel and rapid approaches to identify biomarkers for symptom onset and disease progression to facilitate triage of patients and establish appropriate treatments. Peptidome-based studies using serum from patients and high-throughput spectrometric techniques promise to be valuable for the identification of COVID-19-associated biomarkers. Serum may contain proteins induced by the systemic effects or released to the lung as a result of the viral infection. Thus, patient serum can reflect the physiological or pathological state. Indeed, a proteomic and metabolomic analysis of serum from 46 COVID-19 patients performed by Shen et al demonstrated that using serum proteins and metabolite biomarkers it is possible, not only classify patients according to their grade of severity, but also predict the progression to severe COVID-19 (4). More recently, Messner et al re-designed a high-throughput mass spectrometry platform that enabled the identification of up to 27 potential biomarkers that were differentially expressed depending on the severity grade of COVID-19 (5). Although the technologies used in both studies are highly sensitive and provide robust results, they are time consuming, requires specialized personnel and most importantly, they are not available in most of the hospitals, so their translation bench to bedside is limited.

In the present study, we used MALDI-TOF MS, a simple and fast technology, available in most of the hospitals, to conduct a comparative analysis of serum from 80 COVID-19 patients. Our results demonstrate the value and power of MALDI-TOF to classify and predict the progression of COVID-19 in a clinical setting.

## MATERIALS AND METHODS

### Ethics Statement

The study was conducted in accordance with the Declaration of Helsinki. All human samples were taken after written consent of the participants. They were informed of the purposes of the study, which was approved by the Ethics Review Board of the Illes Balears (CEI).

### Participants

This study included a total of 80 COVID-19 patients who attended Hospital Son Espases, the reference hospital of the Balearic Islands, between March 2020 and June 2020. COVID-19 cases were confirmed based on the Chinese management guideline for COVID-19 (6). Only patients with a positive RT-PCR test were enrolled.

The severity grade of COVID-19 was defined based on the abovementioned guideline (6). Accordingly, COVID-19 patients were classified into three subgroups: mild, severe, or critical. Mild included non-pneumonia and mild pneumonia cases. Severe was characterized by dyspnea, respiratory frequency ≥30/minute, blood oxygen saturation ≤93%, PaO_2_/FiO_2_ ratio <300, and/or lung infiltrates >50% within 24–48 hours. Critical cases were those that exhibited respiratory failure, septic shock, and/or multiple organ dysfunction/failure. Twenty healthy volunteers, including 13 recovered from COVID-19 were also included in the study.

### Sample collection

Blood samples were collected into anticoagulant free-tubes. The tubes were centrifuged at 2,500 rpm at 20°C for 10 min within a 30-min time frame. Serum from each patient sample was then collected, aliquoted and stored at –80°C. Each serum sample was heat inactivated a 56°C for 90 min prior to analysis.

### MALDI-TOF serum sample preparation

The preparation of the serum and the analysis of the samples by MALDI-TOF MS were performed as previously described (7). Serum samples were purified and concentrated using reversed phase C18 tip, Pierce™ C18, following the manufacture instructions. In brief, 20 µl of serum were mixed with 15 µl of trifluoroacetic acid (TFA) 2%. Samples were passed through the Pierce™ C18 tip by pipetting up and down repeatedly (20 times), followed by the separation of the unbound protein solution. After washing the Pierce™ C18 tip with 10 µl of 0.1% TFA, the bound proteins/peptides were eluted with 4 µl of 0.1% TFA:CH_3_CN (1:1, v/v). The solution was passed through the Pierce™ C18 tip repetitively (8-10 times). The eluted protein/peptide solution was mixed with 4 µl of a-cyano-4-hydroxycinnnamic acid (CHCA) matrix solution (10 mg of CHCA in 1 ml of 5% TFA:CH_3_CN, 1:1, v/v) and 1.5 µl of this mixture were spotted onto a MTP 384 target plate ground steel (Bruker Daltonics, Leipzig, Germany) and overlaid with 2 µl of CHCA matrix and allowed to air-drying. The analysis of each sample was conducted in triplicate.

### MALDI-TOF analysis

Measurements were performed on an Autoflex III MALDI-TOF mass spectrometer (Bruker Daltonics, Leipzig, Germany) equipped with a 200 Hz smart beam laser. Spectra were generated by averaging 1000 single laser shots (100 shots at 10 different spot positions) at a laser frequency of 200 Hz and detected in linear positive mode. The IS1 voltage was 20.1 kV, the IS2 voltage was maintained at 18.7 kV, the lens voltage was 8.4 kV, and the extraction delay time was 140 ns. Peaks between 2,000 and 25,400 Da were selected for analysis. Mass accuracy was calibrated externally using the Bruker Bacterial Test Standard. Triplicates of each sample were obtained.

### MALDI-TOF mass data processing

Raw mass spectra obtained by MALDI-TOF MS was analysed using the MALDIquant R package (8). Square root transformation, peak smoothing, baseline correction, and intensity normalization were performed on each mass spectrum. The average spectrum from the triplicates was obtained. Peaks were detected and binned across all average spectra with a signal to noise ratio of 5 and a tolerance of 0.002. Peaks presents in less than 25% of the spectra were rejected. All spectra from the groups under study were pre-processed, and peak detection was applied to obtain an intensity matrix. The resulting matrix of peak intensities was used for Principal Component Analysis (PCA) (9) and Machine learning (ML) approaches (10). To set up ML, the radial basis function was used as the kernel function and ANOVA was used to select the 45 most relevant peaks.

### Liquid chromatography-mass sample preparation

Three microliters of serum were diluted to 1 ml with 50 mM Ammonium bicarbonate (0.2 μg/μl taking the average of plasma proteins as 80 mg/ml). One hundred μL of the dilution were reduced with 11 μl of 50 mM dithiothreitol for 30 minutes at 56°C and were alkylated with 12.5 μl of indole-3-acetic acid 20 mM for 20 minutes in the dark at 37 °C. Total volume 123.5 μl, containing likely 24 μg of plasma protein, were digested with 10 μL of trypsin 100 ng/μl at 37°C overnight. Ten microliters of formic acid (FA) 5% were added to stop the digestion.

### Liquid chromatography-mass setup

The digested peptides were analysed by liquid chromatography mass spectrometry (LC-MS/MS) with a nanoflow Agilent series 1200 LC system (Agilent Technologies, Waldbronn, Germany), with an autosampler equipped with an 8 μl capillary loop, coupled to a Q-Exactive Plus Hybrid Quadrupole-Orbitrap mass spectrometer (ThermoFisher®Scientific) in data dependent acquisition mode. For each acquisition, peptides were loaded onto a precolumn (ZORBAX 300 SB-C18, 5 μm, 5 mm *0.3 mm i.d.) at a flow rate of 15 μl/min for 2 min and then analysed using a 235 min LC gradient (from 3% to 97% buffer B) at a flow rate of 250 nl/min (analytical column, ZORBAX 300 SB-C18, 3.5 μm, 150 mm *0.075 mm i.d.). Buffer A was H_2_O containing FA 0.1%, and buffer B was acetonitrile with 0.1% FA. All reagents were MS grade. The *m/z* range of MS1 was 350-1,650 with the resolution at 140,000 (at 200 m/z), automatic gain control (AGC) target of 3e^6^, and maximum ion injection time (max IT) of 250 ms. Top 10 precursors were selected for MS/MS experiment, with a resolution at 17,500 (at 200 m/z), AGC target of 5e4, and max IT of 200 ms. The isolation window of selected precursor was 4 m/z.

### Liquid chromatography-mass data processing

The resultant mass spectrometric data were analysed using Proteome Discoverer (Version 2.2.0.388, Thermo Fisher Scientific) using a protein database composed of the Homo sapiens fasta database downloaded from UniProtKB on 12 Jul 2020, containing 20,304 reviewed protein sequences, and the SARS-CoV-2 virus fasta downloaded from UniProtKB on 20 May 2020, containing 13 protein sequences. Enzyme was set to trypsin with four missed cleavage tolerance. Static modifications were set to carbamidomethylation (+57.02146) of cysteine and variable modifications were set to oxidation (+15.99492) of methionine and dimethylation (+28.03075) of peptides N-termini. Precursor ion mass tolerance was set to 10 ppm, and product ion mass tolerance was set to 0.6 Da. The peptide-spectrum-match allowed 1% target false discovery rate (FDR) (strict) and 5% target FDR (relaxed). Normalization was performed against the total peptide amount. The other parameters followed the default setup.

## RESULTS

We acquired MALDI mass spectra of 82 serum samples obtained from 80 different COVID-19 patients (from two patients, we analysed two samples (109/144 and 143/141) collected at different severity grades); 34 samples were collected from mild COVID-19 patients, 26 from severe patients and 22 from critical patients. We also analysed 20 serum samples obtained from healthy people, including 13 samples collected from individuals recovered from COVID-19. The relevant characteristics of each group are shown in Table 1.

**Table 1.** Relevant characteristics of the individuals included in this study.

|  | Healthy<br>(n=20) | COVID-19 patients |  |  |
| --- | --- | --- | --- | --- |
|  |  | Mild<br>(n=33) | Severe<br>(n=26) | Critical<br>(n=21) |
| **Sex** |  |  |  |  |
| Male | 11 | 11 | 16 | 21 |
| Female | 9 | 22 | 10 | 0 |
| **Age** |  |  |  |  |
| Mean ± SD | 50±12.3 | 55.2±16.7 | 63.2±9.9 | 63.2±10.1 |
| Median | 50.5 | 49 | 65 | 65.5 |
| Range | 27-73 | 34-89 | 47-81 | 33-76 |

All spectra from the four groups under study were processed and peak detection was applied to obtain an intensity matrix of 135 peaks in the mass range of 2,000 to 25,000 daltons using MALDIquant. To select the most characteristic peaks distinguishing the groups classified according to COVID-19 severity, we applied Post hoc Turkey’s HSD analysis, which identified 78 peaks with a FDR (False Discovery Rate) value ≤ 0.05. Figure 1A illustrates the quantitative variability of those peaks, significantly different, that exhibited a log fold change ≥ 2 for COVID-19 severity on a heatmap. We found clear differences between critical and mild patients. Twelve peaks were significantly different and at least four fold more intense in the set of critical patients than in the mild ones (Figure 1B). However, only two peaks (*m/z* 11,530 and 11,686) were significantly different between severe and critical patients (Figure 1B).

These results encouraged us to apply ML approaches to classify and predict COVID-19 severity. We built two support vector machine learning models using the 135 peaks (*m/z*) obtained from each sample by MALDI-TOF MS. In the first model, patients were classified as severe and non-severe. Severe patients group included those that required oxygen support (severe and critical patients), while non-severe were those without oxygen support (mild patients). Seventy-five percent of the samples from each group were randomly selected to form the training set, while the remaining 25% of the samples were used for validation as an independent test cohort. This process was repeated five independent times, every time ML was tested through fivefold cross-validation. This model reached an average area under curve (AUC) of 0.911 for Receiver Operating Characteristic (ROC) curve (8).

**Figure 1.**
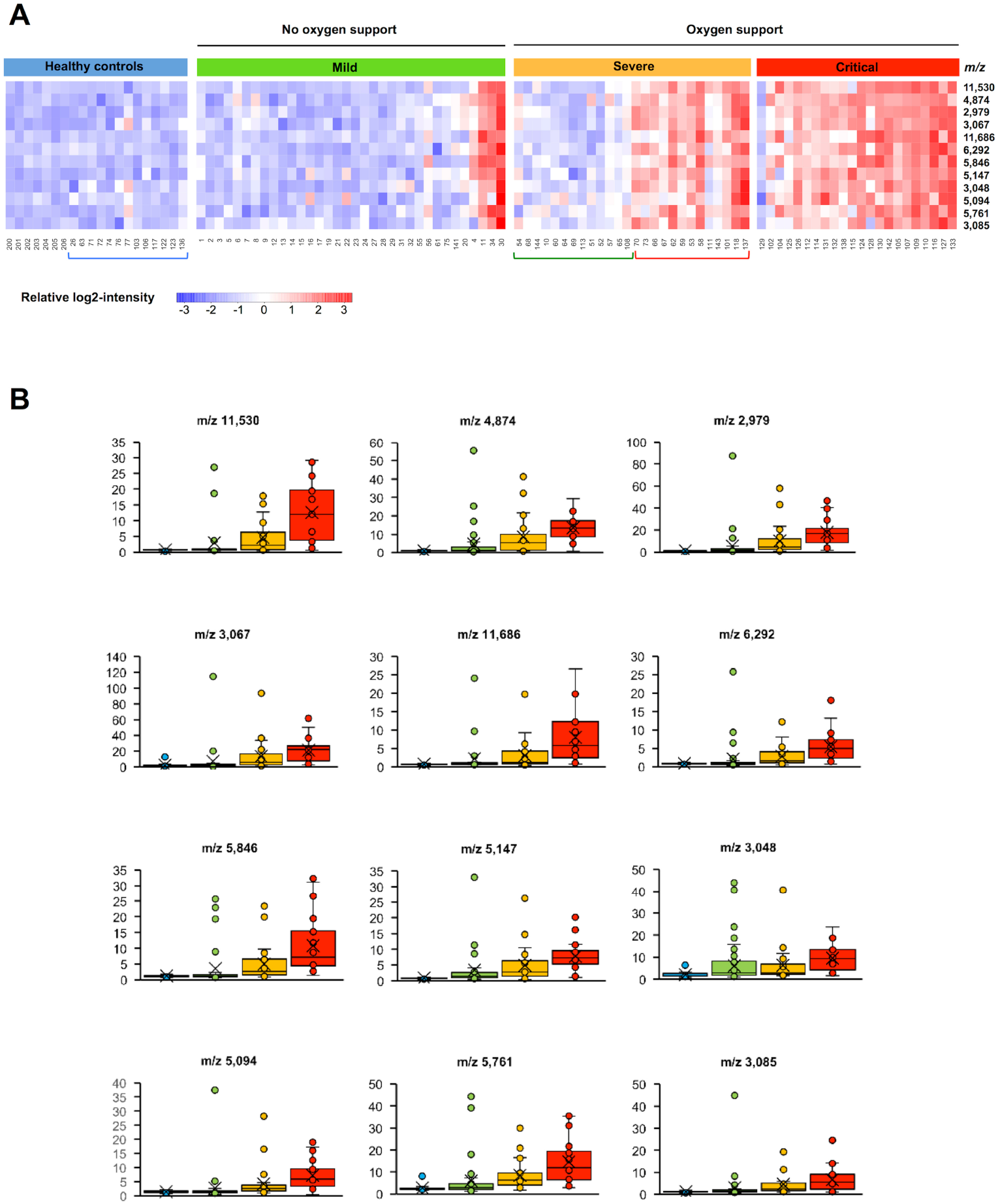
MALDI mass spectra of serum indicate clinical severity in COVID-19. (A) Heatmap illustrates peptidome profiles that inform on COVID-19 severity. Heatmap was generated using the ComplexHeatmap package (8). Groups were classified according to COVID-19 severity following the Chinese management guideline for COVID-19. Blue bracket below heatmap indicates healthy individuals recovered of COVID-19, while green and red brackets indicate samples from patients classified by ML as mild or critical, respectively. (B) Peaks with increased intensity depending on COVID-19 severity. The boxes show the first and third quartiles as well as the median (middle), the mean (cross), and the outliers (circles outside the whiskers).

Using this model, 90 % of the samples were correctly classified with only a 5% of false positives and 5% of false negatives. PCA separated individuals according to the severity of COVID-19 (Figure 2) and helped to identify some outliers (samples 4, 11, 30, 34 from mild patients) previously detected in the heatmap and in the bloxplots.

**Figure 2.**
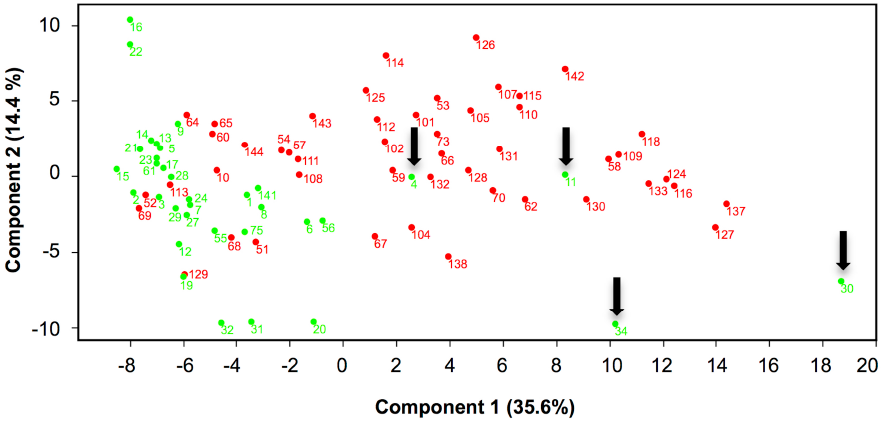
PCA of the mass spectra of the samples from the non-severe (mild) (green dots) and severe (severe and critical) (red dots) patients. Arrows indicate outlier samples.

Next, we assessed whether ML would be able to predict clinical evolution of severe patients (grade 2). For this model, a database constructed using the mass spectra of samples from mild (grade 1) and critical (grade 3) patients, excluding outliers samples, was challenged with the mass spectra of samples from severe patients (grade 2). As above, this process was repeated five independent times, every time ML was tested through fivefold cross-validation. This model reached an AUC of 0.956 for ROC curve. Green and red brackets below heatmap panel in figure 1A indicate those clinically severe (grade 2) patients that were clustered by ML as mild or critical, respectively. All severe patients that were classified by ML as mild had a good clinical evolution except patients of samples 108 and 144, which worsened to critical 8 days later, suggesting that this technology is quite accurate to predict favourable prognostics. On the other hand, five patients (samples 111, 143, 101, 118 and 137) classified as clinically severe (grade 2) when the samples were collected, which 48-96 h later evolved to critical (grade 3), were clearly clustered in the group of critical patients using machine learning, while the remaining 8 patients (70, 73, 66, 67, 62, 59, 53, 58) did not worsen to grade 3.

Most of the discriminating peaks identified by MALDI-TOF MS analysis had a low molecular weight (< 5,000 Da). They probably resulted from the fragmentation of proteins upregulated in severe and critical patients. Interestingly, the most substantial intensity difference were exhibited by the peaks with *m/z* of 11,530 and 11,686, which might correspond to unfragmented proteins of the acute phase induced by the virus. To investigate this hypothesis, we performed a proteomic analysis of the samples by LC MS/MS. We identified five proteins that were significantly upregulated according to the severity of the disease; the serum amyloid A2 protein (SAA2), the C reactive protein (CRP), the serum amyloid protein A1 (SAA1), the lipopolysaccharide binding protein (LBP) and the gamma chain of the fibrinogen (FGG). Figure 3A illustrates the quantitative variability of these proteins on a heatmap. Only the serum level of SAA2 exhibited significant increments between mild and severe patients and between severe and critical patients (Figure 3B). In addition, SAA1, CRP, LBP and FGG were increased in the serum from the critical patients compared with to mild patients, while only CRP and SAA1 were increased in the severe patients compare to mild patients (Figure 3B). Given that the molecular weight of SAA1 and SAA2 is approximately 11.7 kDa, depending on the isoform (11), and that we found a good correlation between the level of both proteins and the intensity of the peaks with *m/z* of 11,530 and 11,686, we suggest that these peaks might correspond to the serum amyloid proteins A1 and A2.

**Figure 3.**
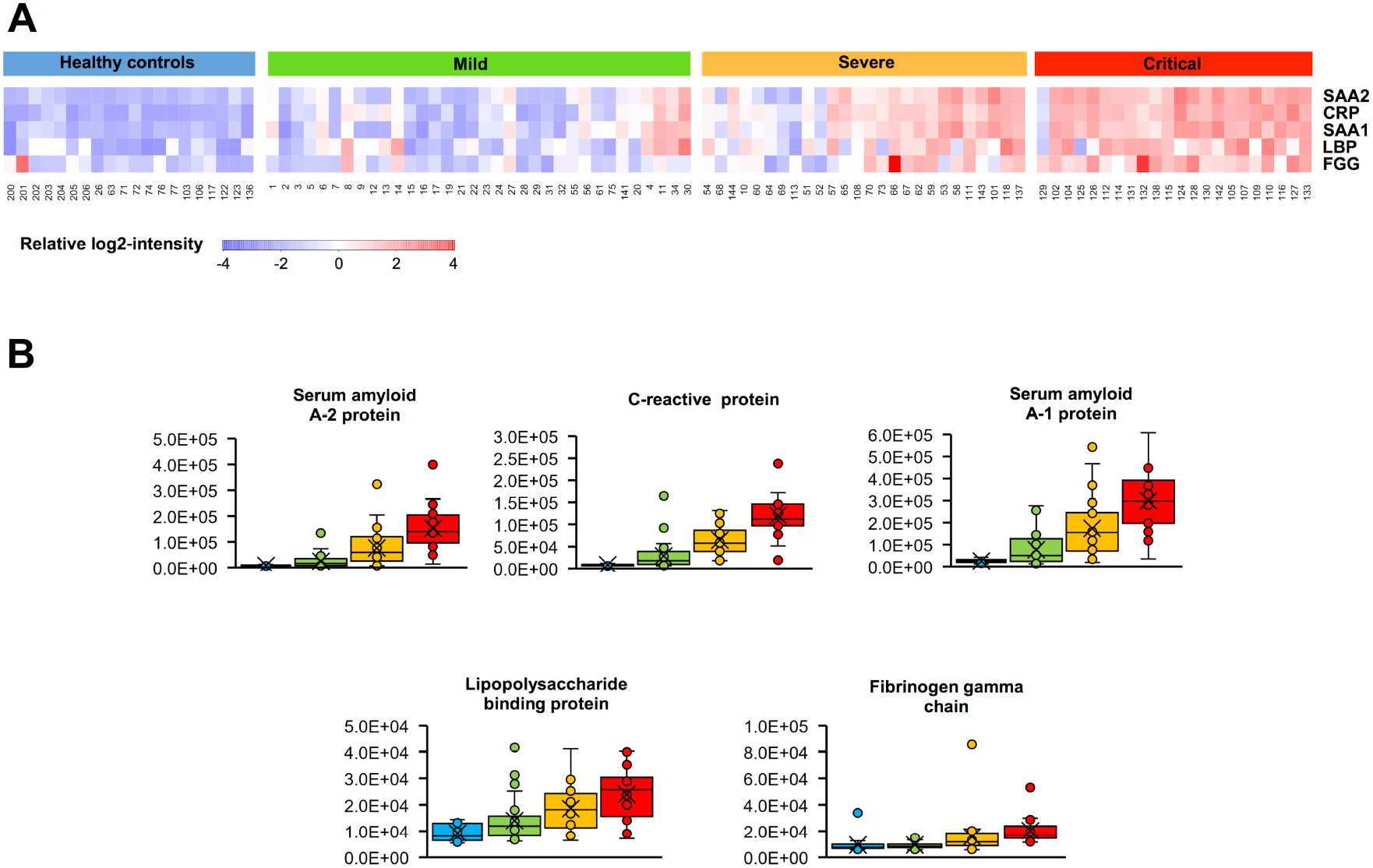
Upregulated proteins according to COVID-19 severity. (A) Heatmap illustrates proteins that inform on COVID-19 severity. Heatmap was generated using the ComplexHeatmap package (8). Only proteins that were present in more than 70% of the samples identified by at least 5 peptides and significantly increased (P< 0.05) by a log-2 fold change >2 were included in the figure. Groups were classified according to COVID-19 severity following the Chinese management guideline for COVID-19. SAA2 (serum amyloid A2 protein), CRP (C reactive protein), SAA1 (serum amyloid protein A1), LBP (lipopolysaccharide binding protein) and FGG (gamma chain of the fibrinogen). (B) Upregulated proteins depending on COVID-19 severity. The boxes show the first and third quartiles as well as the median (middle), the mean (cross), and the outliers (circles outside the whiskers).

## DISCUSSION

In this study we demonstrate that the molecular changes that occur in the sera of COVID-19 patients may be detected by MALDI-TOF MS analysis generating peptidome profiles that may be used as clinical classifiers. In addition, we show that it is possible to predict the progression of the disease using the peptidome signatures obtained with this technology. Finally, we provide strong evidences that serum amyloid A2 protein is one of the major biomarkers of severe COVID-19 disease.

To our knowledge, only two previous studies reported the use of mass spectrometry analysis of serum from COVID-19 patients to classify disease severity (4, 5). However, both studies were performed using sophisticated technologies, which are not available in most of the hospitals. Our challenge was to test whether MALDI-TOF MS analysis, a simpler technology available in most of the clinical microbiology laboratories for identification of microbial species, was able to achieve similar results. Our study classified severe patients with a high accuracy (90%), very similar to that obtained in a previous report (93%) (4), with a very low number of false results.

We represented the changes of those peaks that exhibited major changes upon grouping the patients according to CDC Chinese severity criteria, ranging from scale 1 to scale 3 in a heatmap, which graphically illustrated how level changes in these peaks reflected a progression from mild to critical COVID-19. Interestingly, our peptidome profile data identify the most important changes within the severe patients, upon which a patient is put on oxygen supply. This observation is consistent with the proteome analysis conducted by Messner et al, who found that at molecular level the requirement of oxygen supply coincided with the progression to severe disease (5). In contrast, mild patients have a peptidome signature virtually identical to the healthy controls suggesting that in non-severe patients changes are restricted to the site of infection, the respiratory tract, without significant molecular systemic alterations. Bloxplot representation also identified outlier samples that were confirmed by PCA. As occurred in the study by Messner et al. (5), it is likely that these samples were collected from patients with certain underlying pathologies o under specific treatments that altered the peptidome signature.

Case studies demonstrated the clinical utility of the peptidome profiles to classify and predict COVID-19 evolution. First, five samples from patients classified as clinically severe, presented a profile very similar to the group of critical patients in the heatmap and clustered in this group using ML 48-96 h before they clinically progressed to critical. Second, sample 141 obtained from the same patient as sample 143, which evolved from severe to critical, was collected two days before it was discharged and presented a peptidome profile clustered as mild by ML and in the heatmap. On the other hand, samples from patients completely recovered of COVID-19 (blue bracket in figure 1A) or those that evolved from severe to mild had peptidome signatures similar to those observed in the healthy control patients. Conversely, peptidome profile of sample 144, obtained from the same patient as sample 109, but eight days before he progressed to critical, was not able to predict the clinical trajectory of this patient.

Overall, these results suggest that peptidome profiles obtained by MALDI-TOF MS may represent a good prognostic tool to support clinical decisions. However, it would be interesting to conduct a longitudinal study with sequential daily samples from a cohort of patients at different grades of severity until their recovery to assess the anticipation time of prediction.

One of the potential limitations of our study is that due to the rapid response required in the initial stages of the pandemic situation, we collected samples from the patients that were admitted in our hospital using as unique criteria that they were hospitalized due to a SARS-CoV-2 infection. Therefore our study did not take in account some confounding factors, like age. Nonetheless, the change of the intensity of the peaks between groups substantially exceeded the variability observed within each group with ages ranging from 33 to 89, suggesting that differences in the peptidomes profiles of different groups are poorly influenced by confounding factors.

Two of the most intense peaks detected in the sera from critical patients had *m/z* of 11,530 and 11,686 that might correspond to two different isoforms of the serum amyloid A protein (11). This acute phase markers, induced by the proinflammatory cytokine IL-6, were two of the predominant proteins detected by both Shen et al and Messner et al in their respective studies (4,5). As in our study, they also detected CRP, LBP and FGG as clear protein biomarkers for COVID-19 severity.

In conclusion, our study supports the potential of the MALDI-TOF MS as a fast and clinically available technology to aid clinicians in their decisions on COVID-19 patients and identifies serum amyloid protein A2 as an excellent biomarker to monitor COVID-19 patients.

## Data Availability

All Data are available

## ACKNOWLEDGMENTS

The authors thank Sara Fernández for helpful discussions on statistical analysis. This study was funded by Instituto de Investigación Sanitaria de las Islas Baleares (IdISBa).

